# An all-inclusive electrical impedance tomography system for remote disease monitoring of multiple vital organs

**DOI:** 10.64898/2026.04.27.26351830

**Authors:** James H.W. Li, Bradley J. Edelman, Wang C. Kwok, Michael Lawson, Ravi Bahukhandi, Henry Lui, Iris Y. Zhou, Kevin C. Chan, Kai Hang Yiu, Erik Fung, Russell W. Chan

## Abstract

Chronic cardiopulmonary, metabolic, and renal diseases represent an immense global health burden, yet access to organ-specific diagnostics remains limited outside of hospitals. Most clinical assessments rely on imaging or laboratory testing that is costly, infrastructure-dependent, and impractical for large-scale or longitudinal monitoring in community settings. Here, we introduce VitoCheck, a compact, user-friendly electrical impedance tomography (EIT) platform that provides non-invasive evaluation of lung, heart, liver, and kidney function within minutes. We first demonstrate system stability, spatial specificity, and spectral sensitivity through controlled phantom studies. We then validate VitoCheck in clinical cohorts by demonstrating accurate EIT-based predictions of standard diagnostic metrics, including spirometry-derived forced expiratory volumes, echocardiography-derived ejection fraction, ultrasound-derived liver fat scores, and blood serum-derived kidney filtration. User feedback further highlights the rapid scan workflow that supports deployment by non-specialists in decentralized environments. By combining portable and easy-to-use hardware with quantitative organ health analytics, VitoCheck enables scalable screening and proactive disease management for use in remote and out-of-clinic care.

## Introduction

Chronic diseases of the lungs, heart, liver, and kidneys are among leading causes of morbidity, mortality, and healthcare expenditure worldwide^1^. Conditions such as chronic obstructive pulmonary disease (COPD), ischemic heart disease and cardiomyopathy, metabolic dysfunction-associated steatotic liver disease (MASLD) and cirrhosis, and chronic kidney disease (CKD) affect billions worldwide and contribute substantially to disability, hospitalization, and mortality^2–5^. Since early-stage dysfunction is often asymptomatic and diagnoses delayed until advanced organ damage and complications have already occurred^6^, these missed opportunities narrow the window for preventive intervention and increase long-term clinical burden and healthcare costs.^7,8^ Current diagnostic strategies rely heavily on tools such as computed tomography (CT), magnetic resonance imaging (MRI), ultrasound, and laboratory assays. While clinically informative, these modalities are not point-of-care and require costly infrastructure, specialized training, and hospital workflows^9,10^. Moreover, access to these technologies remains limited in rural and low-resource settings where specialist availability and diagnostic capacity frequently lag behind disease prevalence^11^. Even within well-resourced healthcare systems, logistical constraints such as specialist waitlists or frequent travel to centralized facilities can impede routine monitoring for individuals with chronic or progressive diseases. As health systems increasingly prioritize decentralized and preventive models of care, there is growing need for portable technologies that are capable of providing organ-specific assessments independent of centralized imaging or laboratory resources^9,12^. A user-friendly solution easily deployable in community settings could improve chronic disease management while easing the strain on healthcare infrastructure^13,14^.

Electrical impedance tomography (EIT) offers a promising basis for such technology. EIT uses low-voltage surface electrodes to reconstruct conductivity distributions linked to air, fluid, and soft tissue properties^15^. Unlike most traditional clinical imaging systems, EIT systems are radiation-free, portable, and relatively resilient to operator error, making them suitable for continuous use in non-hospital environments^16,17^. Despite these attractive qualities, existing EIT systems have remained largely restricted to pulmonary monitoring during ventilation procedures in the ICU^18,19^. Nevertheless, additional studies have shown that EIT can independently capture physiological signatures of regional ventilation, cardiac dynamics, hepatic fat content, and kidney tissue composition^20–23^. These works suggest that EIT can serve as a multipurpose tool for monitoring chronic organ dysfunction. However, few studies and frameworks have demonstrated that EIT can scale reliably outside hospital environments where minimally trained operators must scan diverse populations and provide on-demand diagnostic results.

To address this need, we introduce VitoCheck, a compact EIT platform designed for decentralized, multi-organ health assessment. Rapid and device-guided data acquisition protocols, combined with on-the-fly machine learning-based analytics pipelines transform impedance measurements into quantitative surrogates for lung function, cardiac contractility, liver steatosis, and kidney filtration to provide clinically interpretable metrics linked to chronic disease states. By delivering an all-inclusive digital organ health assessment, VitoCheck aims to expand access to organ-specific diagnostics in settings where health data are commonly limited to basic anthropometric measures and where access to imaging or laboratory testing is scarce^24^. Such environments include population health screenings, primary-care offices, community health fairs, nursing homes, rural clinics, and more. Moreover, multi-organ capability is particularly advantageous given the clinical co-occurrence of cardiopulmonary, metabolic, and renal disorders in aging populations and in individuals with metabolic risk factors^25,26^.

In this work, we present (i) the design and characterization of the VitoCheck system, (ii) validation in controlled phantom experiments to assess sensitivity to spatial and spectral conductivity changes, and (iii) *in vivo* evaluation across lung, heart, liver, and kidney assessments with benchmarking against standard clinical measures. We further evaluate device usability, operator training requirements, scan duration, and participant acceptance to assess its practicality for decentralized deployment. Together, these results demonstrate a portable EIT platform capable of multi-organ assessment outside traditional clinical environments.

## Results

### VitoCheck system overview and characterization

The VitoCheck system comprises a handheld console and a flexible 16-electrode belt containing fully integrated current-drive and voltage-measurement electronics that enable a rapid self-guided setup and high-quality data acquisition (**Fig. 1a-c**). The belt can be positioned at organ-specific anatomical landmarks (**Fig. 1d**), which allows the same hardware to interrogate lung, heart, liver, and kidney function with only slight modification to its physical location. A core requirement for reliable multi-organ EIT is the generation of stable voltage measurements across all electrode configurations. VitoCheck uses a pair of drive electrodes (source/sink) and a pair of measurement electrodes for each configuration (**Fig. 1e**), yielding 208 unique measurement pairs per excitation frequency. Representative raw voltage data (“U-curves”) depicting amplitude across electrode measurement pairs exhibit the expected periodic peaks with minimal noise contamination (**Fig. 1f**). When condensed as a function of electrode spacing, the mean U-curve demonstrates the expected rapid decay in amplitude with increasing circular distance between drive and measurement electrode pairs (**Fig. 1g**). These behaviors reaffirm proper current injection and electrode contact, and facilitate the detection of dirty datasets that do not fit such canonical patterns (**Fig. S1a-c).**

**Fig. 1.**
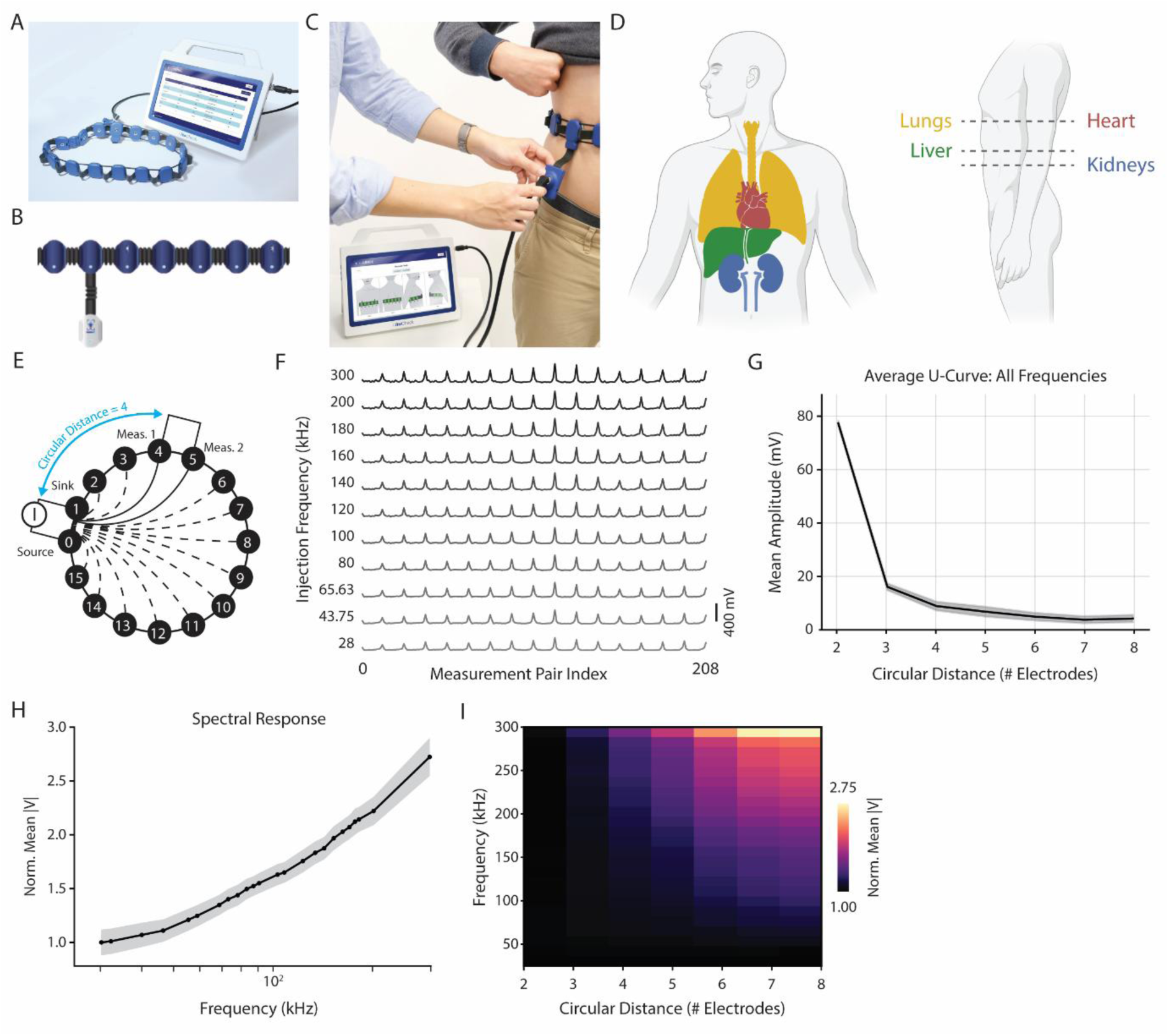
Overview and characterization of the VitoCheck portable EIT system. **(A)** The VitoCheck platform consists of a handheld console and a flexible 16-electrode belt for rapid multi-frequency electrical impedance tomography (EIT). **(B)** Close-up of the electrode belt showing the integrated electronics module that performs current injection and voltage sensing. **(C)** Example clinical application showing belt placement around the abdomen. The console guides correct placement, verifies electrode contact quality, and initiates automated data acquisition. **(D)** Anatomical overview of the four organs (lungs, heart, liver, and kidneys) assessed with VitoCheck and the corresponding belt positions. **(E)** Illustration of the rotational measurement protocol using adjacent current injection and the corresponding measurement electrodes. Circular electrode distance is defined as the number of electrodes separating a given measurement pair from the drive pair. **(F)** Representative multi-frequency U-curves (28 - 300 kHz) demonstrating stable, periodic amplitude patterns across all 208 electrode-pair measurements and frequencies. **(G)** Average U-curve (pooled across frequencies) showing the expected monotonic decay in amplitude with increasing circular electrode distance. **(H)** Normalized spectral response of the system from a representative dataset, computed as the mean voltage across all measurement pairs, relative to the lowest injection frequency. **(I)** Heatmap of the normalized spectral response across circular distances, demonstrating consistent frequency-dependent amplitude scaling and stable behavior across measurement geometry. Shaded regions indicate standard error on the mean.

Beyond spatial consistency, we also assessed VitoCheck’s spectral performance by measuring the normalized mean voltage across 15 frequencies between 28 kHz and 300 kHz. The resulting spectral response (**Fig. 1h**) captures the expected monotonic increase in admittance with frequency and is consistent across datasets (**Fig. S1d**)^27^. Mapping this frequency-dependent behavior across circular electrode distance further highlights the system’s stability and sensitivity to tissue impedance where higher frequencies exhibit greater voltage spread and interelectrode coupling (**Fig. 1i, Fig. S1e**).

### Phantom validation using time-difference and frequency-difference EIT

Because high-quality voltage data are a prerequisite for accurate EIT imaging, we next examined whether VitoCheck’s measurement stability enables consistent reconstruction of conductivity changes (**Fig. 2a)**. To do so, we designed various phantom experiments that simulate physiological conditions such as fluid shifts, airflow differences, and tissue composition abnormalities^28^, and assessed these changes using either time-difference EIT (tdEIT)^29^ or frequency-difference EIT (fdEIT)^30^ approaches (**Fig. 2b**).

**Fig. 2.**
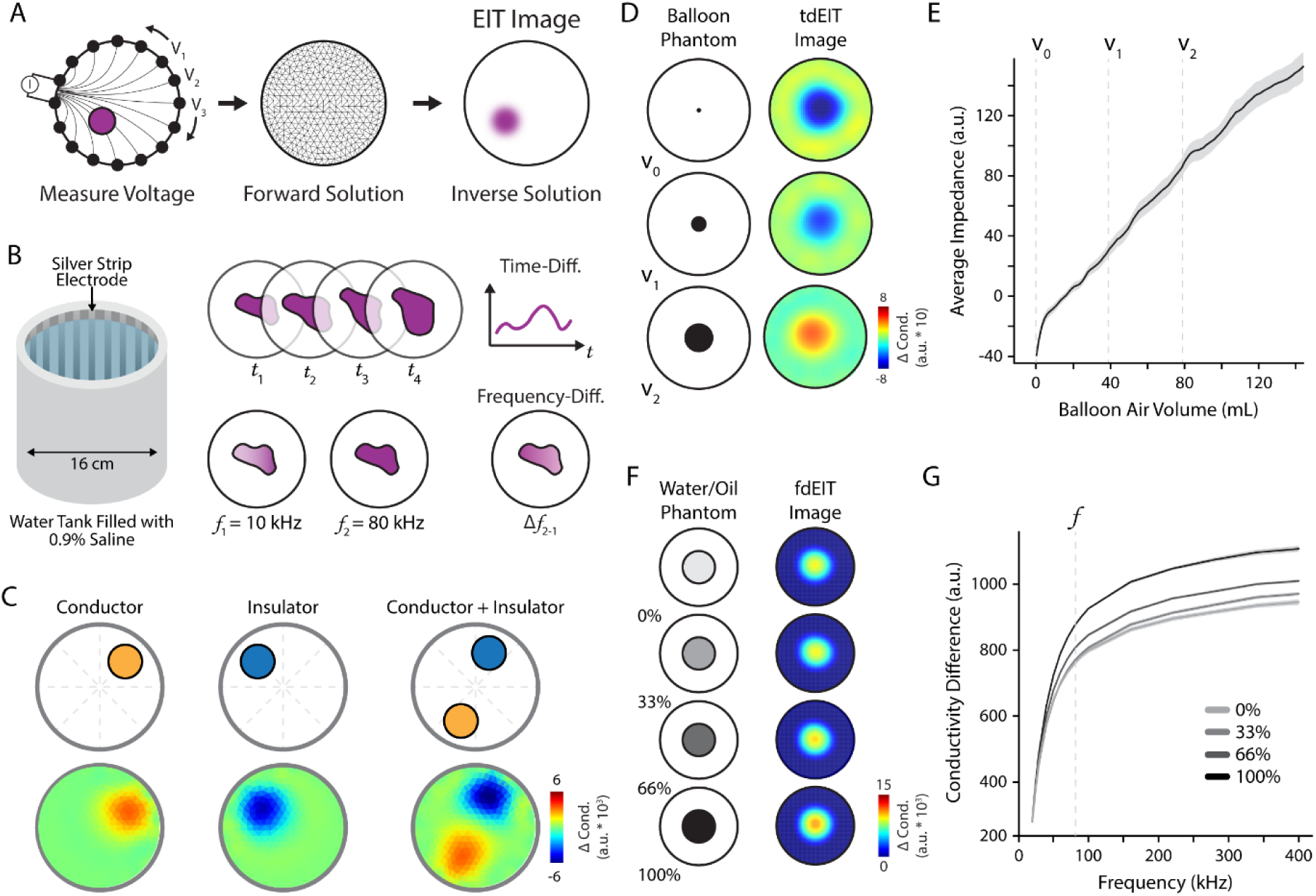
Phantom validation of VitoCheck demonstrating *in vitro* time- and frequency-difference imaging. **(A)** Electrical impedance tomography (EIT) comprises measuring surface voltages and then reconstructing internal conductivity images via forward and inverse solutions. **(B)** Phantom setup with saline-filled tank and silver perimeter electrodes. Time-difference EIT (tdEIT) compares frames over time and frequency-difference EIT (fdEIT) compares responses at different frequencies. **(C)** Visual representation of the placement of acrylic (insulator) and aluminum (conductor) inclusions. The tdEIT reconstructions reveal accurate localization and polarity for both inclusion types. **(D)** tdEIT reconstructions of an increasing balloon phantom (representing non-conductive air volumes) reveal a corresponding impedance displacement. **(E)** The average impedance is linearly related to theoretical balloon volume. **(F)** Oil-water phantoms with increasing conductive concentrations and the corresponding fdEIT reconstructions (80 kHz, 10 kHz reference). **(G)** Frequency response plot from 20 to 400 kHz (10 kHz reference) of reconstructed conductivity differences across phantom conductive concentrations. Shaded regions represent standard deviation across measurements. Two-way ANOVA with main effects of frequency and oil-water proportion, ***p < 0.001.

We first introduced inclusions with contrasting electrical properties to a homogeneous saline-filled tank (**Fig. S2a)**. Reconstructed images revealed spatially localized impedance changes corresponding to each physical inclusion, with inverse polarities reflecting the opposing conductivities (**Fig. 2c**). Next, to simulate dynamic physiological changes such as volume shifts in the lungs, we incrementally inflated an air-filled balloon submerged in saline and reconstructed images at a fixed frame rate (**Fig. 2d, Fig. S2b**). Resulting tdEIT images captured the evolving spatial impedance distribution as the balloon displaced the surrounding fluid. Quantitative analysis revealed a monotonic and linear increase in reconstructed impedance magnitude with balloon volume (R^2^ = 0.99, p < 0.001) (**Fig. 2e**). These results demonstrate the system’s ability to detect and distinguish both conductive and insulating anomalies relative to a known baseline, and confirm its ability to capture gradual geometric alterations.

We further assessed VitoCheck’s fdEIT capabilities using varying concentrations of oil-water mixtures to simulate tissues with varying adiposity. Image reconstructions comparing impedance responses revealed consistent differences across solutions with increasing conductivity enhancements (**Fig. 2f, Fig. S2c**). The spectral response plot also revealed a clear and significant separation of impedance magnitudes across the range of excitation frequencies, indicating that the system can detect material-specific conductivity differences even in the absence of geometric variation (**Fig. 2g, Fig. S2d, Table S1-2**). These findings further demonstrate that VitoCheck is capable of both spatial localization and spectral discrimination.

### Multi-organ health monitoring aligns with clinical reference standards

To ensure that these reconstruction capabilities translate reliably to human imaging where motion and improper belt placement can introduce noise, we developed a machine learning-guided quality-control (QC) framework for the automated detection and correction of faulty electrodes and corrupted frames (**Fig. S3-S4**). During data acquisition, each electrode participates in current injection and voltage measurement in four distinct roles for each frame: (1) current source, (2) current sink, (3) measurement 1, and (4) measurement 2. We therefore engineered four role-specific amplitude features per electrode (**Fig. S3a-b**), which exhibit stable frequency-dependent trends in clean data but show characteristic distortions under poor contact conditions (**Fig. S3c-d**). These features feed into a pre-trained random forest classifier that labels each electrode, frame, or frequency as either clean or faulty (**Fig. S4a-b**).

Across large collections of datasets, this framework enables fine-grained identification of both transient contact loss and frequency-specific corruption, producing intuitive summaries of frame- and electrode-level data quality (**Fig. S4c-f**). Electrodes or frequencies labelled as faulty are interpolated in the temporal (tdEIT) or spectral (fdEIT) domain, respectively, ensuring stable forward-model inputs before image reconstruction. Although the phantom experiments showed minimal corruption due to their controlled conditions, this automated QC pipeline is essential for human imaging, where it provides fully unsupervised filtering of correctable or unusable measurements.

With this QC layer in place, we next applied VitoCheck to *in vivo* multi-organ assessment and validated organ-specific analysis pipelines designed for the lungs, heart, liver, and kidneys (**Fig. S5**)^22,31–33^. For pulmonary assessment, tdEIT was used to quantify ventilation dynamics during forced breathing maneuvers. As expected, individuals with impaired lung function exhibited markedly slower volume rise compared to healthy controls, particularly during the first half of the maneuver (**Fig. 3a-b, Table S3**). Corresponding EIT images revealed diminished regional ventilation and increased spatial heterogeneity throughout the breathing cycle (**Fig. 3c-f, Fig. S6a-b, Table S4**). Importantly, increased heterogeneity was restricted to the right lung, consistent with its greater susceptibility to airflow obstruction due to its larger volume (three lobes vs. two) and steeper main bronchus.

**Fig. 3.**
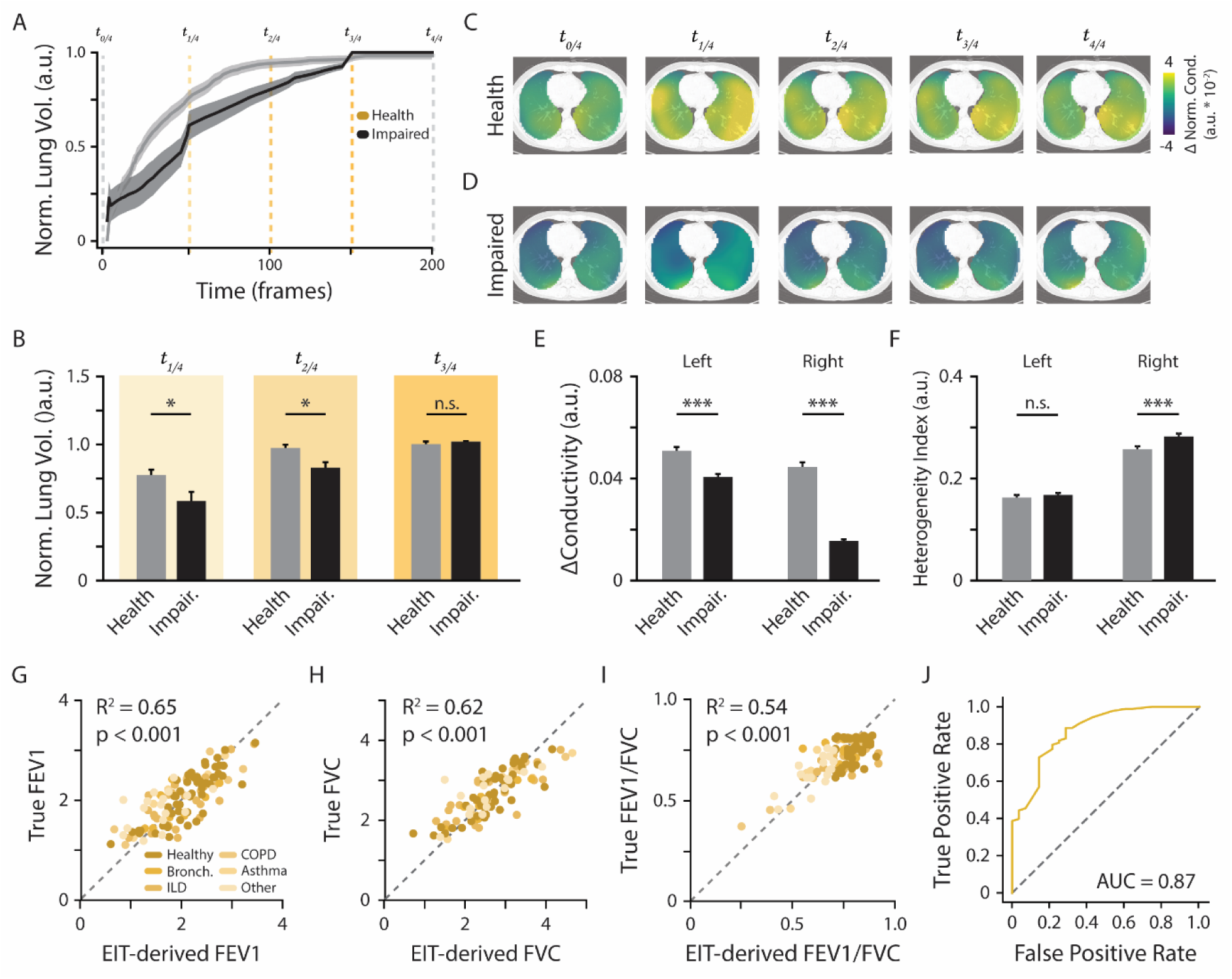
Clinical validation of VitoCheck for lung assessment. **(A)** Normalized forced-breathing curves for individuals with healthy (grey) and impaired (black) lung function. Shaded regions denote inter-subject standard deviation. **(B)** Group-level comparison of normalized lung volume at time points t_1/4_ - t_3/4_ shows significantly reduced expansion in impaired individuals at early and mid-procedure time points. Welch’s t-test, *p < 0.05, false-discovery rate corrected; n.s. = not significant. **(C-D)** Time-resolved EIT images at five time points of the forced breathing procedure (t_0/4_ - t_4/4_) for healthy participants (C) and those with lung impairments (D). **(E)** Quantification of conductivity change in the left and right lungs reveals significant reductions in impaired lung patients compared to healthy individuals. **(D)** Heterogeneity index analysis shows significantly greater spatial ventilation asymmetry in the right impaired lung compared to the right healthy lung. For (E) and (F), Unpaired t-test, ***p < 0.001, false-discovery rate corrected; n.s. = not significant. Bars represent the mean, and error bars indicate standard error on the mean. **(G-I)** Scatterplots comparing EIT-derived predictions with spirometry reference values demonstrate strong correlations for FEV_1_ (R^2^ = 0.65), FVC (R^2^ = 0.62), and the Tiffeneau–Pinelli Index (FEV_1/_FVC; R^2^ = 0.54); all p < 0.001. **(J)** Receiver operating characteristic curve for the classification of impaired vs healthy lung function based on EIT-derived FEV_1/_FVC, achieving an AUC of 0.87.

To evaluate whether VitoCheck can serve as a surrogate for spirometry, tdEIT-derived features were combined with anthropometric parameters to predict standard lung metrics. EIT predictions strongly correlated with clinically measured forced expiratory volume (FEV_1_; R^2^ = 0.65, p < 0.001), forced vital capacity (FVC; R^2^ = 0.62, p < 0.001), and the Tiffeneau-Pinelli ratio (FEV_1_/FVC; R^2^ = 0.54, p < 0.001) (**Fig. 3g-i**). Further examination of feature importance based on SHapley Additive exPlanations (SHAP) showed that conductivity-derived measures contributed substantially alongside anthropometric variables (**Fig. S6d-f**). This indicates that EIT provides complementary physiological information rather than merely reproducing trends captured by bodily measurements. Finally, VitoCheck-based classification of obstructive impairment achieved a receiver operating characteristic (ROC) area under the curve (AUC) of 0.87, with a sensitivity of 83% and specificity of 73% (**Fig. 3j**). Together, these results demonstrate that VitoCheck can quantify ventilation mechanics and capture clinically relevant spatial abnormalities of lung function.

EIT-based cardiac assessment was performed using a 3-lead ECG-gated approach to estimate left ventricular ejection fraction (LVEF). In the current cohort, raw ECG traces revealed reduced end-diastolic and end-systolic amplitudes in individuals with echocardiography-defined impaired cardiac function (**Fig. 4a-c, Fig. S7a-b, Table S5**), however, the difference between end-diastole and end-systole was not sufficient to distinguish groups (**Fig. 4d**). To extract and compare impedance-based counterparts, tdEIT frames were synchronized to R-peaks and T-waves to represent end-diastole and end-systole, respectively. Conductivity amplitudes within a cardiac region-of-interest showed visibly reduced end-diastolic and end-systolic magnitudes in individuals with low LVEF compared to those with healthy LVEF (**Fig. 4e**). Quantitative analysis confirmed this effect, revealing significant differences between groups at both time points and in the overall end-diastole to end-systole change (**Fig. 4f-h, Table S5**). This added discriminatory power suggests that EIT detects ventricular motion and blood-volume changes that a 3-lead ECG does not reliably capture. As a result, impedance-based monitoring may provide stronger discrimination where reliable, but more complex 12-lead ECG are not available^34^.

**Fig. 4.**
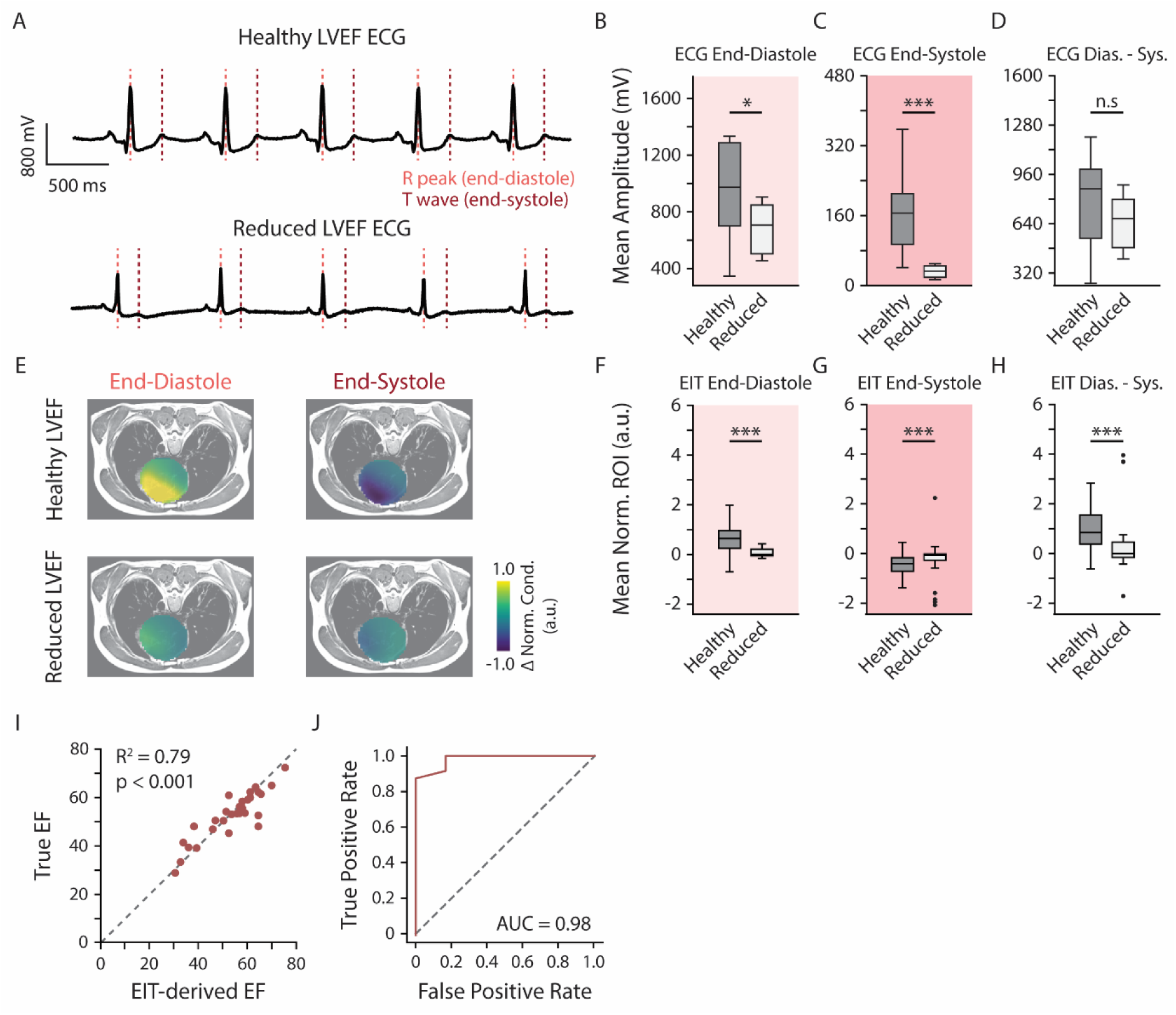
Clinical validation of VitoCheck for cardiac function assessment. **(A)** Representative electrocardiogram (ECG) traces from individuals with healthy versus reduced left ventricular ejection fraction (LVEF). Red dashed lines denote automated detection of the R-peak (end-diastole) and T-wave (end-systole), used for synchronizing EIT signals to the cardiac cycle. **(B-D)** Group-level ECG amplitude features showing reduced end-diastolic and end-systolic peak amplitudes in reduced-LVEF cases, while end-diastole to end-systole differences were not significantly different. **(E)** Group-level end-diastolic and end-systolic EIT cardiac images. Individuals with healthy LVEF show strong contractile-associated conductivity changes, whereas those with reduced LVEF demonstrate blunted systolic response and reduced diastolic recovery. (**F-H**) Quantification of EIT-derived cardiac features reveals significantly reduced end-diastolic and end-systolic regional conductivity changes and a diminished end-diastole to end-systole contrast in participants with reduced LVEF. For (B-D) and (F-H), Welch’s t-test, *p < 0.05, ***p < 0.005; n.s. = not statistically significant. Box plots represent median ± interquartile range. **(I)** EIT-derived and echocardiography-measured LVEF were strongly correlated (R^2^ = 0.79, p < 0.001). **(J)** Receiver operating characteristic curve analysis shows excellent discrimination between normal and reduced LVEF (AUC = 0.98).

For classification, engineered features were based on segmented myocardial regions fit to an ellipse model (**Fig. S7c**), which enabled quantification of contraction volume, spatial heterogeneity, and the relative size of end-diastolic and end-systolic conductivity changes. Using such features, EIT-derived estimates of LVEF closely matched echocardiographic reference measurements (R^2^ = 0.79, p < 0.001) (**Fig. 4i**) and enabled accurate classification of individuals with reduced LVEF (AUC = 0.98, sensitivity = 83%, specificity = 96%) (**Fig. 4j, Fig. S7d)**). Model interpretability using SHAP scores revealed that the relative size of EIT-derived R-peak and T-wave regions and contraction velocity contributed most strongly to predictions (**Fig. S7e**), reflecting physiologically relevant markers of myocardial weakness. Together, these findings show that ECG-gated EIT enables non-invasive quantification of cardiac function.

To assess hepatic steatosis, we evaluated frequency-difference EIT reconstructions in MASLD and healthy states. CAP scores derived from VitoCheck closely matched ultrasound-based clinical measurements (R^2^ = 0.67, p < 0.001) (**Fig. 5a**), and EIT-derived features reliably classified MASLD with high discriminative performance (AUC = 0.95, sensitivity = 88%, specificity = 92%) (**Fig. 5b, Fig. S8a**). Across all models, SHAP analysis revealed that the most influential predictors were broadband frequency-difference features, with diverse spectral pairs consistently driving MASLD classification across subjects (**Fig. S8c-e**).

**Fig. 5.**
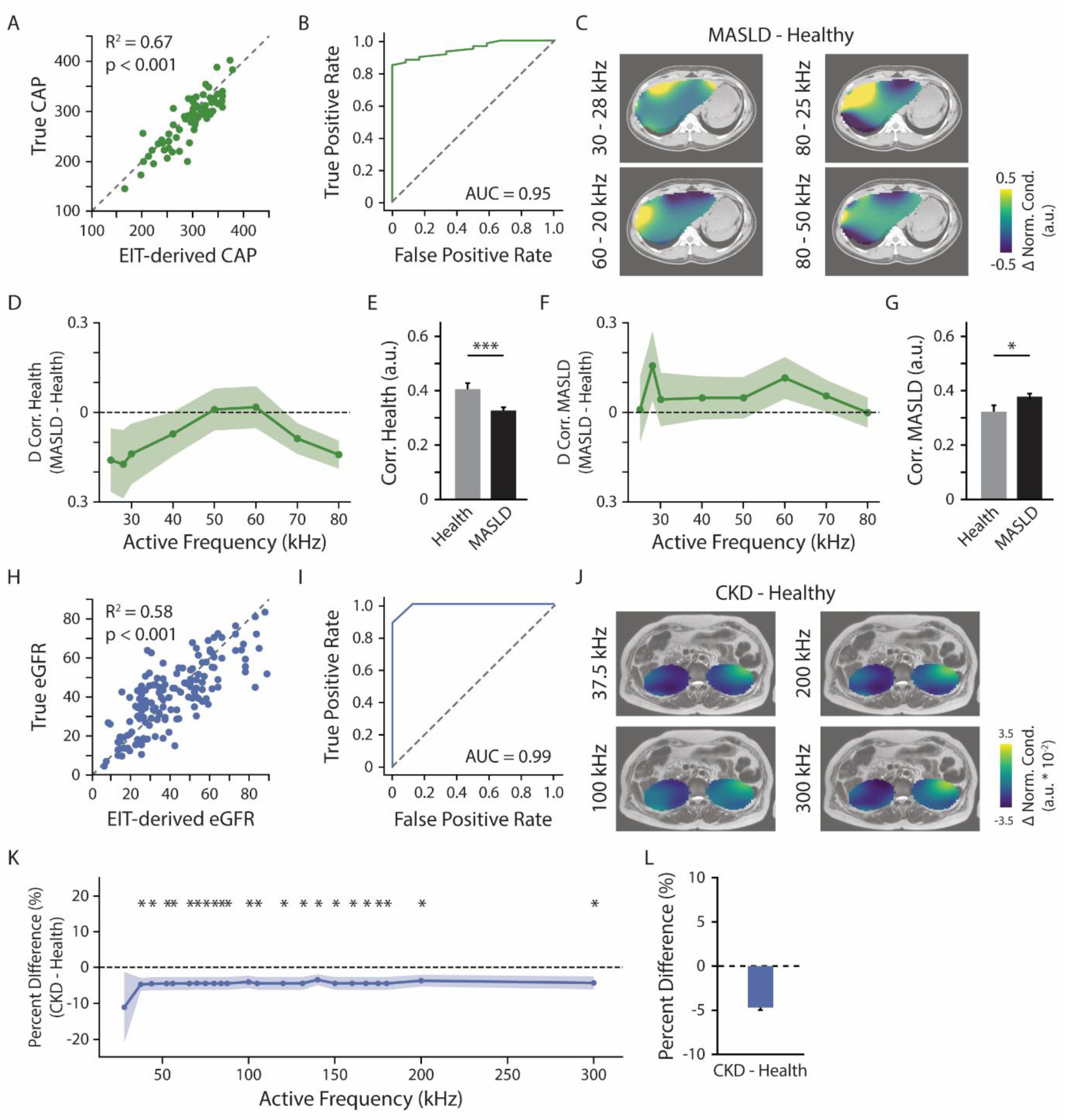
Liver and kidney assessment using spectral-spatial EIT features. **(A)** EIT-derived controlled attenuation parameter (CAP) scores strongly correlated with ultrasound-based clinical CAP measurements (R^2^ = 0.67, p < 0.001). **(B)** Receiver operating characteristic (ROC) curve demonstrating high discriminative performance for classifying MASLD and healthy states using EIT-derived features (AUC = 0.95). **(C)** Group-level frequency-difference reconstructions showing distinct spectral-spatial conductivity patterns in MASLD compared to healthy liver across varying frequency pairs. **(D-E)** Spatial correlation of individual liver maps to healthy templates revealed reduced similarity in MASLD compared to the healthy state across excitation frequencies (D). Aggregate similarity values demonstrate that MASLD maps were significantly less correlated to healthy templates (E). **(F-G)** Complementary analysis using MASLD templates demonstrated increased correlation in MASLD subjects relative to healthy participants (F), confirming greater similarity to steatotic liver signatures (G). For (D) and (F), two-way ANOVA with main effects of excitation frequency and disease state: *p < 0.05, ***p < 0.005 **(H)** Regression analysis showing strong agreement between EIT-derived estimated glomerular filtration rate (eGFR) and clinical laboratory values (R^2^ = 0.58, p < 0.001). **(I)** ROC curve demonstrating high discriminative performance of EIT-based models in identifying CKD, achieving an AUC of 0.99. **(J)** Representative frequency-difference EIT images highlighting spatial conductivity differences between CKD and healthy states. **(K)** Group-level kidney ROI percent difference for CKD - healthy state across all excitation frequencies. Welch’s t-test, *p < 0.05, false-discovery rate corrected; n.s. = not significant. **(L)** Broadband summary of the effect size (percent difference) across the full frequency spectrum. Bars represent the mean, and error bars indicate standard error on the mean.

Spectral contrast maps revealed distinct spatial conductivity responses in MASLD across frequency pairs (**Fig. 5c**, **Fig. S8b**), which motivated a template-matching strategy to quantify these patterns. For each fdEIT pair, group-average maps were generated to produce a healthy and MASLD template. Individual liver images were then compared against these references using pixel-wise spatial correlation. MASLD images showed significantly reduced similarity to the healthy template across most active frequencies (**Fig. 5d-e, Table S6**), whereas correlation to disease templates was consistently higher than healthy images (**Fig. 5f-g, Table S7**). This complementary analysis demonstrates that MASLD is characterized by frequency-dependent spatial patterns of impedance change that systematically deviate from healthy liver structure. The reciprocal increase in the disease template similarity confirms that MASLD images share reproducible signatures that can be detected non-invasively. Together, these results show that VitoCheck can capture and quantify both the magnitude and topology of frequency-dependent liver health.

EIT measurements of renal function also strongly correlated with eGFR calculated from blood serum collections (R^2^ = 0.58, p < 0.001) (**Fig. 5h**). The VitoCheck system also effectively differentiated healthy kidney function from CKD status (AUC = 0.99, sensitivity = 99%, specificity = 100%) (**Fig. 5i, Fig. S9a**). Notably, age emerged as the highest-ranking feature in the SHAP analysis, which is consistent with its well-established role as the strongest epidemiological risk factor for CKD (**Fig. S9b**)^35^. However, several principal component-derived features also captured spatial information that contributed to distinguishing disease state (**Fig. S9c**).

Similar to liver imaging, fdEIT images highlight spatial conductivity differences between healthy and CKD states that may represent renal perfusion or fluid status (**Fig. 5j, Fig. S9c**). Nevertheless, in contrast to the liver, these kidney maps did not exhibit overtly distinct spatial patterns across frequencies. Therefore, we focused on determining whether the magnitude of these differences varied systematically with excitation frequency. When assessing the percent difference in conductivity in a bilateral kidney ROI between CKD and healthy states, we found a robust and consistent reduction in CKD across nearly all frequencies, with significant group differences spanning most of the spectral range (**Fig. 5k, Table S8**). Despite these consistent differences, a mixed-effects model did not detect a global main effect of group (**Table S9**), consistent with the modest effect size relative to within-subject variability.

### EIT improves diagnostic value and acceptance in non-hospital settings

Finally, we evaluated whether VitoCheck can provide actionable diagnostic value in settings where clinical infrastructure and diagnostic expertise are limited. In many community and primary-care environments, anthropometric measurements are often the only available health indicators, which may exhibit limited physiological specificity for the detection of early organ dysfunction. By providing quantitative surrogates for clinical diagnostics, VitoCheck enables proactive disease management outside of traditional clinical settings. To assess this benefit, we compared EIT-enhanced models (as described in previous sections) against anthropometric-only baselines for identifying lung dysfunction, cardiac contractile impairment, liver steatosis risk, and chronic kidney disease (**Fig. 6a-d**). Across all four organ systems, EIT-derived models showed superior classification performance (AUC_EIT_ = 0.87 - 0.99) relative to anthropometric-only models (AUC_ANTHROP_ = 0.69 - 0.90), demonstrating that EIT captures physiological information that is inaccessible to size- or age-based screening alone.

**Fig. 6.**
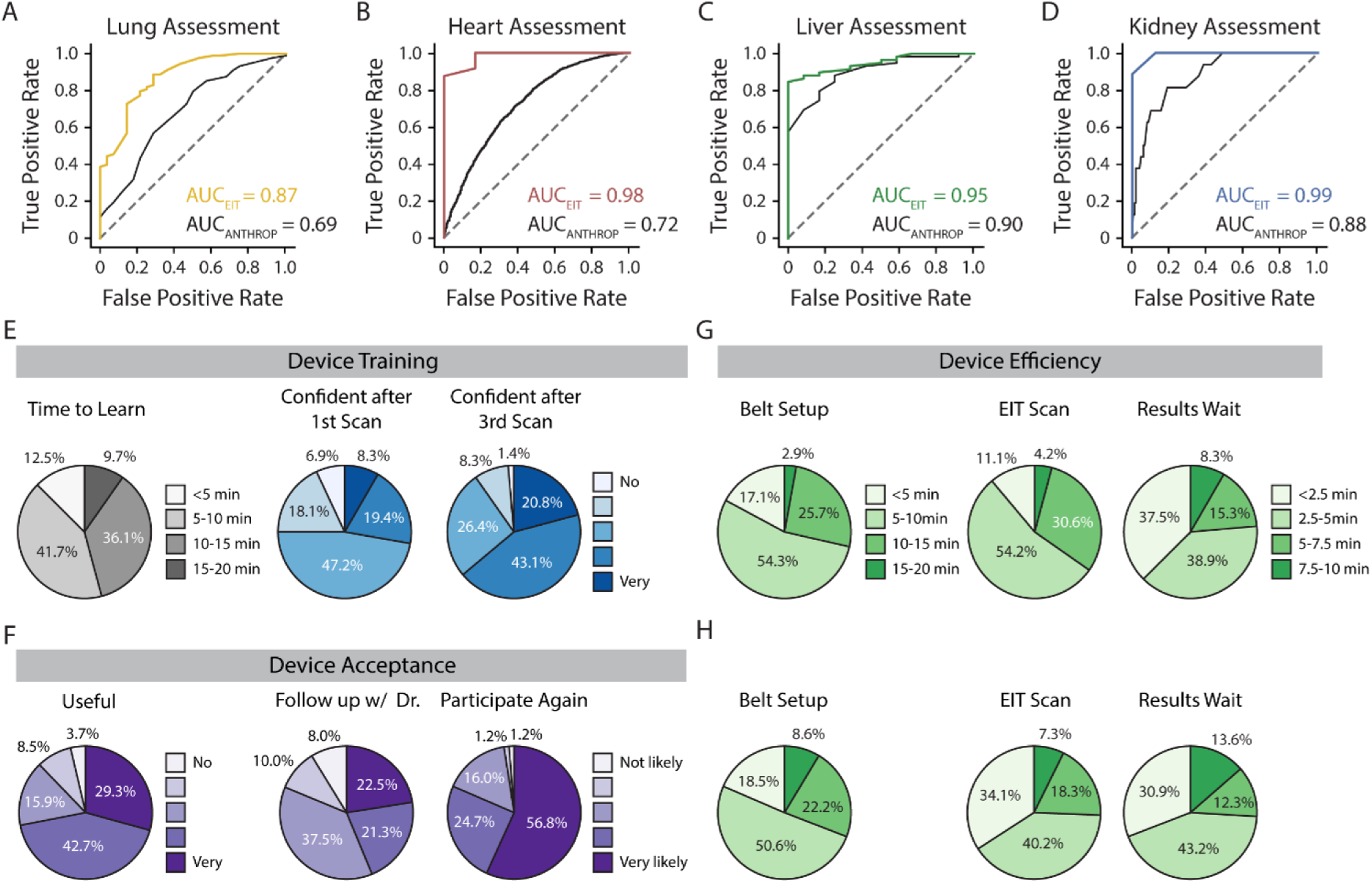
VitoCheck subjectively and objectively. **(A-D)** Receiver operating characteristic curves comparing disease-state classification using EIT-derived features (colored lines) versus anthropometric-only baseline models (black). EIT substantially improved discrimination for (A) impaired lung function (AUC_EIT_ = 0.87 vs. AUC_ANTHROP_ = 0.69), (B) reduced left ventricular ejection fraction (AUC_EIT_ = 0.98 vs. AUC_ANTHROP_ = 0.72), (C) MASLD (AUC_EIT_ = 0.95 vs. AUC_ANTHROP_ = 0.90), and (D) CKD (AUC_EIT_ = 0.99 vs. AUC_ANTHROP_ = 0.88). **(E-H)** Operator and participant surveys demonstrating system accessibility, efficiency, and acceptance during community screening. **(E)** Operators overwhelmingly reported training times <15 min (90.3%) and rapid improvement in confidence after the first and third scans (confident or very confident: first 27.7%, third 63.9%). **(F)** Participants found the device useful or very useful (72.0%), were likely to follow up with a clinician (43.8%), and strongly indicated a willingness to participate in future screenings (81.5%). **(G-H)** Reported times for belt setup were consistently under 10 minutes for both operators (71.4%) and participants (69.1%). Reported times for EIT scanning and results delivery were under 5 minutes for both administrators (65.3% scanning, 76.4% results) (G) and participants (74.3% scanning, 74.1% results) (H), highlighting the feasibility of rapid, high-throughput deployment in non-hospital environments.

Importantly, these advances were achieved with a rapid and highly usable workflow that support the deployment of VitoCheck by non-experts. Training surveys (n = 73) showed that after only 15 minutes and three scans, nearly 65% of operators felt confident in using the device, a rapid improvement from just over 25% of operators after one scan (**Fig. 6e**). Operators further found the device easy to use, highly portable, and useful for community screenings, with 86% recommending its use in non-hospital settings (**Fig. S10a-d**). Such usability aligns with growing evidence that community health workers can reliably perform non-invasive screening when supported by simple diagnostic tools^36^. Participant survey feedback (n = 83) further highlighted excellent comfort, understanding, and acceptance of the VitoCheck scan with 99% willing to undergo a repeat scan and 86% recommending the test to others (**Fig. S11a-e**). Moreover, participants reported favorable scores on the utility of the VitoCheck scan, especially compared to other clinical technologies, and found that the results both matched their expectations and motivated them to follow up with a doctor (**Fig. 6f, Fig. S11f-g**). Finally, when juxtaposing the reported efficiency of the VitoCheck device, the majority of operators and participants reported that the entire process (i.e., belt setup, EIT scan, and result delivery) was completed within 15 minutes (**Fig. 6g-h**). Together, these results establish VitoCheck as a practical, scalable, and patient-friendly screening solution. By delivering real-time organ-function biomarkers beyond what anthropometric measurements provide, VitoCheck extends diagnostic capabilities into underserved communities where existing tools cannot reliably detect early disease.

## Discussion

Together, our results establish VitoCheck as a robust, portable platform capable of delivering clinically relevant multi-organ information using EIT outside of traditional hospital environments. By addressing limitations in electrode stability VitoCheck promotes high-fidelity impedance imaging in community settings where noise, motion, and operator variability have historically hindered EIT adoption. Another key feature that facilitates signal stability is a reference electrode that reduces variability, especially in healthy individuals, and which provides more robust baseline measurements to aid in the detection disease states (**Fig. S12a**)^37^. When combined with embedded analytics pipelines, QC safeguards and improved signal stability ensure that reliable physiological data can be obtained by non-expert operators, which is essential for real-world application and scalability. In addition, preliminary exploration of reduced electrode setups, combined with artificial intelligence strategies to preserve image quality, shows promise for further enhancing system usability and flexibility in non-expert environments (**Fig. S12b-c**).

Applications for this device include pulmonary function tracking, cardiac monitoring, liver fat assessment, and renal function estimation. By leveraging time-difference and frequency-difference EIT, the system captures a diverse set of physiological signatures ranging from ventilation heterogeneity, end-diastolic and end-systolic conductivity changes, steatotic spectral patterns, to kidney-specific conductivity deficits that align with clinical reference standards. These metrics enable diagnostic-grade quantification of dysfunction across multiple organs, democratizing the early detection of diseases in the community that otherwise often remain latent until advanced stages. Simultaneous multi-organ monitoring also provides a unique advantage in assessing comorbidities such as COPD, cardiomyopathy, and heart failure, where overlapping symptoms complicate diagnosis^38^. In such cases, concurrent imaging of cardiopulmonary dynamics (**Fig. S13a-c**) may enable more holistic, personalized assessment and treatment planning. Similarly, liver and kidney dysfunction frequently co-occur in metabolic and chronic disease states^39^, which may reflect shared systemic disease pathways that also overlap with diabetes mellitus and atherosclerotic cardiovascular diseases. The close anatomical proximity of these organs may allow EIT measurements to be obtained with minimal or no adjustment of the electrode belt during a single screening session. The modular and flexible nature of this EIT platform may also be extended to future applications in gastrointestinal dysfunction^40^, neurovascular disorders such as stroke^41^, or even cancer detection via dielectric profiling^42^.

Compared with health assessments relying primarily on questionnaires, anthropometric measurements^43^, and simple physiological recordings such as blood pressure^44^ or temperature^45^, VitoCheck-derived metrics capture physiologically actionable signatures including regional ventilation heterogeneity, LVEF, and frequency-dependent liver and kidney impedance patterns that align with established clinical reference standards across organ systems. Importantly, these EIT-based features provide discriminative power beyond demographic and anthropometric variables alone, as reflected by improved classification performance across all organ assessments. This suggests that VitoCheck does not merely detect manifest disease but also captures underlying alterations that may precede traditional clinical diagnosis. Overall, our findings demonstrate the feasibility of EIT imaging from controlled *in vitro* imaging to *in vivo* screening of multi-organ diseases. Its non-invasive and portable design supports deployment in remote care frameworks to improve access for patients in resource-limited environments, providing a decentralized alternative to safeguard the health of a larger community. The system also holds promise for population screening for important chronic medical illnesses, in which early detection will enable timely and effective treatments to prevent organ failure.

## Methods

### VitoCheck EIT System

VitoCheck (E-SENSE Innovation & Technology Ltd, Hong Kong) is a portable EIT platform consisting of a 16-electrode belt, a reference electrode, and a console interface. For phantom imaging, VitoCheck was attached to a 0.9% saline-filled tank (Diameter = 16 cm) with evenly spaced silver electrode strips placed on the wall. Signals were acquired by connecting the acquisition module and the console interface to the electrodes using alligator clips. For human imaging, the electrode belt was placed around the appropriate anatomical region. For lung and heart imaging, the belt was placed at the level of the nipples for men and just below the breasts for women. For liver and kidney imaging, the belt was placed just above or below the lowest rib, respectively. Gel-based patches ensured contact between the skin and electrodes; contact impedance was measured and visualized during setup to ensure proper contact between the electrodes and the skin.

### Phantom EIT Data Acquisition

Three sets of phantom experiments were performed to assess the ability to (1) detect opposing conductivity polarities, (2) detect geometric variations in conductivity profiles, and (3) detect variations in conductivity profiles. For the first set of experiments, acrylic (insulator) and aluminum (conductor) cylinders were placed at different locations within the saline-filled tank. For the second set of experiments, a latex balloon was placed at the center of the tank and was inflated at a constant rate of 26 mL/s for 6 s. For both of these experiments, EIT measurements were acquired at an excitation frequency of 100 kHz at a frame rate of 33 Hz. Measurements were averaged over a 3 s period for the static insulator/conductor imaging. For the third set of experiments, pure peanut oil and distilled water were mixed with two teaspoons (10 mL) of detergent at concentrations of 0%, 33%, 66% and 100%. For this experiment, EIT measurements were acquired at 17 excitation frequencies between 10 kHz and 400 kHz (i.e. 10 kHz, 20 kHz, 25 kHz, 28 kHz, 30 kHz, 40 kHz, 50 kHz, 60 kHz, 70 kHz, 80 kHz, 90 kHz, 100 kHz, 160 kHz, 220 kHz, 280 kHz, 340 kHz, 400 kHz). Three replicates were performed for the first and second experiments, and five times for the third experiment.

### Phantom EIT Image Reconstruction and Analysis

All EIT images were reconstructed using a Gauss-Newton solver with Newton’s One-Step Error Reconstruction (NOSER) regularization from the pyEIT toolbox^46^. Time-difference EIT (tdEIT) reconstruction for balloon phantom experiment was performed using the signal average as reference. Frequency-difference EIT (fdEIT) reconstruction for oil-water experiments was performed using measurements at different pairs of frequencies. Images were then transformed into a 64 x 64 grid using interpolation coefficients defined by a sigmoid function. The average value at a fixed region of interest (ROI) for the balloon and oil-water phantom was extracted.

### Clinical Study Participants

The lung study enrolled 116 participants (age 20 - 86 years, 44 male), including 50 healthy controls and 66 patients (interstitial lung disease: 26, bronchiectasis: 9, asthma: 8, COPD: 6, others: 17). Participants with unstable heart or lung conditions and those unable to perform spirometry (i.e. unable to follow instructions, with end-tracheostomy etc.) were excluded.

The cardiac study enrolled 30 participants (age 22 - 64 years, 16 male), including 15 healthy controls and 15 heart failure patients. Participants with previously documented heart disease (for healthy controls) or any heart disease other than heart failure (for patients), as well as those who had a previous heart or lung surgery or transplantation were excluded.

The liver study enrolled 71 participants (age 19 - 76 years, 30 male), including 12 healthy controls and 59 patients with clinically confirmed MASLD. Participants with previous liver disease (for healthy controls) or any liver disease other than MASLD (for patients) were excluded. Additional exclusion criteria included previous liver surgery or transplantation, ascites, or heavy alcohol intake.

The kidney study enrolled 194 participants (age 26 - 93 years, 110 male), including 18 healthy controls and 176 patients with clinically confirmed CKD. Participants with recent kidney or abdominal surgery were excluded.

For all studies, participants were excluded if they had a recent chest or abdominal surgery, damaged skin on the chest or abdomen, implanted electronic or metallic devices, spinal diseases/discomfort, or if they were pregnant.

### Informed Consent

Informed Consent was provided to recruited participants across four studies targeting kidney, lung, heart, and liver function. All protocols were approved by the Institutional Review Board of The University of Hong Kong (lung: UW-21-579, kidney: UW-21-579, liver: UW-21-265) or the Hospital Authority Hong Kong West Cluster (heart: HKSTP-CREC 2024-023)

### Anthropometric Data Collection

Anthropometric information including height, weight, age, ethnicity, and biological sex was collected from study participants. Chest circumference was obtained from participants in the lung and heart study, whereas waist circumference was obtained from participants of liver and kidney trials. Participants also answered a series of questions including but not limited to alcohol use, exercise habits, and history of familial liver and heart disease.

### Clinical Reference Standards

Lung function was assessed using standard spirometry (CareFusion/Beckton Dickinson, San Diego, CA, USA), by trained operators according to standard clinical practice. During spirometry, study participants were instructed to wear the electrode belt during established clinical spirometry protocols. For this procedure, participants were instructed to take a deep, full inhalation, followed by a forceful exhalation for at least 6 s until reaching a volume plateau indicated by the spirometer. Three replicates were performed, and the best trial was chosen by trained operators. COPD diagnosis, or more generally impaired lung function, was determined based on an FEV_1_/FVC ratio < 0.7 according to the Global Initiative for Obstructive Lung Disease (GOLD) criteria^47^.

The eGFR value was calculated using the CKD-EPI creatinine equation^48^ based on each study participant’s serum creatinine levels. Healthy CKD values were defined as those > 90 mL/min/1.73m^49^. eGFR greater than 90 was annotated as ‘> 90’ due to eGFR values being less reliable above this threshold level^50^. Other eGFR values < 90 remained numerical for regression and classification purposes.

Liver CAP scores were measured using FibroScan (Echosens, Paris, France) by trained operators according to standard clinical practice. Participants were instructed to fast for at least 4 hours prior to the scan. Healthy CAP scores were defined as those ≤ 248 (dB/m)^51^.

LVEF of the heart was obtained through standard echocardiographic measurements acquired using the Resona 7 ultrasound system (Mindray, Shenzhen, China). Risk LVEF values were defined as those < 45%^52^.

### Clinical EIT Data Acquisition

Study participants were instructed to remain still and breathe normally (kidney/heart) or to follow screen-prompted spirometry (lung) or breath-hold (liver) instructions. Time-resolved EIT data was acquired at 33 Hz for lung, kidney and liver measurements and at 107 Hz for heart measurements. The sampling rate of the heart EIT recordings was maximized (according to device capabilities) to align with simultaneous three-lead ECG acquired using an integrated chip within the VitoCheck system. The increased sampling rate is necessary to capture the rapid dynamics of the R-peaks and T-waves of the ECG signal that represent ventricular diastole and systole, respectively, and which are used as temporal landmarks for calculating ejection fraction.

Lung EIT was acquired using an excitation frequency of 100 kHz, simultaneously with spirometry measurements according to the standard protocol previously described in the *Clinical Reference Standards* section. Cardiac EIT measurements consisted of 1-3 one min of spontaneous breathing using an excitation frequency of 100 kHz. A total of 24 excitation frequencies between 28 kHz and 300 kHz (i.e. 28 kHz, 30 kHz, 37.5 kHz, 43.75 kHz, 52.5 kHz, 56 kHz, 65.625 kHz, 70 kHz, 75 kHz, 80 kHz, 84 kHz, 87.5 kHz, 100 kHz, 105 kHz, 120 kHz, 131.25 kHz, 140 kHz, 150 kHz, 160 kHz, 168 kHz, 175 kHz, 180 kHz, 200 kHz, 300 kHz) were used for kidney imaging, and consisted of three segments of 1-min recordings. For liver imaging, 10 frequencies between 20 kHz and 100 kHz (i.e. 20 kHz, 25 kHz, 30 kHz, 40 kHz, 50 kHz, 60 kHz, 70 kHz, 80 kHz, 90 kHz, 100 kHz) were used. Liver data acquisition consisted of 5 frequency profile recordings lasting 5-7 s each (breath-hold) for a total of 25 - 35 s, performed in triplicate.

### EIT Image Reconstruction

Raw EIT data was first cleaned using a machine learning-aided faulty electrode detection pipeline. A random forest classifier was trained using both clean data and data with known faulty electrodes. Faulty electrode data were generated by an operator physically detaching individual electrodes from multiple participants during data acquisition, which enabled the training data to be clearly labeled as either clean or faulty. For each frame, four features from each of the 16 electrodes were combined to create a feature vector of length 1 x 64. The features included the sum of signal intensity when the electrode acted as the (1) current source, (2) current sink, (3) first voltage measurement electrode, (4) second voltage measurement electrode. The classifier then determined for each frame whether faulty electrodes were present. For tdEIT, individual faulty electrodes were linearly interpolated in time, and for fdEIT whole frames containing one or more faulty electrodes were discarded. For fdEIT, missing frequency measurements or those with less than 5% clean frames were interpolated in the spectral domain using polynomial interpolation.

Similar to the phantom studies, EIT images were reconstructed using a Gauss-Newton solver with NOSER regularization from the pyEIT toolbox^46^. Liver prior constructed from CT image was utilized during liver fdEIT reconstruction (weight = 0.01) while prior-less reconstruction was used for other pipelines. Images were then transformed into a 64 x 64 grid using interpolation coefficients defined by a sigmoid function for easier extraction of useful features. tdEIT was applied to lung and heart data at specific time points/windows for further analyses, as described in subsequent sections. fdEIT was applied to liver and kidney data by extracting difference images between pairs of frequencies, with one acting as the reference and the other as the frequency of interest.

### Lung EIT Feature Extraction

For lung EIT, force-breathing curves were extracted from the transformed tdEIT images using uniform manifold approximation and projection (UMAP). Briefly, after preprocessing, tdEIT data were temporally smoothed using a moving average filter and then projected onto a single latent UMAP dimension, preserving dynamic breathing patterns. This one-dimensional representation was used to align breathing cycles across individuals by detecting regions of high temporal variance corresponding to inhalation and exhalation phases. Maximum gradients and extrapolated 1-s gradients were then extracted as the FVC and FEV_1_ metrics, respectively. For the FEV_1_/FVC model, the predicted FEV_1_/FVC ratio based on the predicted FVC and FEV_1_values from the individual models, was also included. Anthropometric data such as age, height, weight, chest circumference, sex, ethnicity, and BMI were included as additional features.

### Regional Lung Metrics

Regional conductivity values were calculated using the maximum difference of images interpolated by the UMAP-generated breathing curve. The conductivity map was then generated using the difference between the highest measured lung volume against the lowest.

### Heart EIT Feature Extraction

EIT data were used to derive quantitative hemodynamic and electrophysiological indicators of heart function. To do so, ECG data were first filtered using a 3rd order low-pass Butterworth filter to remove baseline drift. R-peaks and T-waves were then identified within the clean time-series using BioSPPy, an open source biosignal processing toolbox^53^. The EIT images corresponding to R-peaks and the end of T-waves were averaged to represent impedance distributions at end-diastole and end-systole, respectively. A heart region of interest (ROI) was derived by identifying the area exhibiting the largest impedance change between averaged and normalized end-diastole and end-systole images. The average heart ROI value in these frames, as well as the ratio between the two were used as features.

An 80% threshold was applied to the average images based on the maximum value of the ROI in each image, and an elliptical contour was fitted to the largest continuous region remaining (smaller regions representing spatial noise were ignored). These contours represent left ventricular volume at end-diastole and end-systole, and were used as additional features.

Other additional features included contraction velocity, spatial heterogeneity, and repolarization gradient. The contraction velocity, 𝑉_𝑐_, reflects the spatial gradient of myocardial shortening, with higher values indicating coordinated ventricular contraction, and was derived as follows:

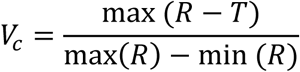

Where max (𝑅 − 𝑇) represents the maximum instantaneous difference between depolarisation (contraction-phase, R) and repolarisation (relaxation-phase, T) conductivity values, normalized by the dynamic range of the depolarisation frame.

Spatial heterogeneity was calculated during both depolarization, 𝜎_𝑅_, and repolarisation, 𝜎_𝑇_, to assess regional desynchrony, and was computed as follows:

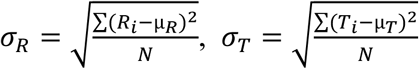

Where µ_𝑅_ and µ_𝑇_ are the mean conductivity values during depolarization, R, and repolarization, T, respectively, with N the number of pixels in the ventricular ROI.

The repolarization dispersion, 𝒢_𝑇_ gradient provides a measure of ventricular filling and stroke volume, and identifies regions with abrupt repolarisation timing shifts, which mechanically manifest as delayed diastolic relaxation This feature was defined as the maximum spatial gradient of conductivity values during the repolarization phase, as follows:

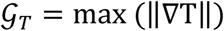

Where ∇T is the spatial gradient of T.

### Liver EIT Feature Extraction

For liver data, fdEIT images were created using various pairwise combinations of excitation frequencies, resulting in 55 images. Each map was processed using a modified ResNet-18 pretrained on ImageNet, in which the first convolutional layer was adapted to accept single-channel EIT images. The 512-dimensional feature embedding prior to the classification head was extracted for downstream learning, yielding 28,160 deep features per subject. In parallel, voxel intensities from fdEIT reconstructions at nine discrete frequencies (20, 25, 28, 30, 40, 50, 60, 70, 80 kHz) were flattened and concatenated as handcrafted features. Treating each voxel as an independent feature produced 323,072 total features when combined with the ResNet embeddings. Anthropometric data such as age, height, weight, waist circumference, and sex were included as additional features.

### Kidney EIT Feature Extraction

For kidney data, the reference frequency was set to 30 kHz, resulting in 23 fdEIT images. The fdEIT images from each individual were then pooled together and subjected to PCA to identify kidney-related components. Conductivity-related features were then extracted from such components to represent the average conductivity value inside a defined kidney ROI, outside a defined kidney ROI, and inside the entire field of view. Anthropometric data such as age, height, weight, waist circumference, sex, and BMI were included as additional features.

### Regression Analysis

Ensemble learning regression models were used to predict organ health metrics from EIT-derived counterparts. Random forest regression was applied to lung metrics and extreme gradient boosting (XGBoost) regression was applied to heart and kidney metrics. Hyperparameter tuning was implemented via 3- or 5-fold cross validation to identify optimal model parameters. After parameter selection, model performance was re-evaluated using an additional round of 3- or 5-fold cross-validation. For liver metrics, a 3-fold elastic net regression model was first trained to predict a CAP baseline score using waist-to-height ratio, body height, and the two most predictive ResNet-derived embeddings. To refine residual error, an XGBoost model was subsequently trained to predict the difference between the ground-truth and elastic-net-estimated CAP scores. This residual learner used 67 additional ResNet image features, 12 voxel-intensity features, and waist-to-height ratio (80 total features). Final CAP predictions were obtained by summing the elastic-net and XGBoost outputs, and the mean of all model instances (3 elastic-net + 3 XGBoost models) minus 12 was used as the final subject-level estimate. When appropriate, class-weighting was implemented to account for imbalanced class structure. The receiver operating characteristic (ROC) and regression plots from the highest-performing fold are reported.

### Receiver Operating Characteristic (ROC) Analysis

For all organ systems, ROC curves were generated to assess the binary classification performance of EIT + anthropometric data-derived models in distinguishing healthy versus diseased organ states, as defined in the *Clinical Reference Standards* section. The area-under-the-ROC curve (AUC) was used as a summary metric of model discriminative power, with values closer to 1.0 indicating higher classification accuracy. Confusion matrices were computed to quantify classification outcomes at the clinically defined thresholds, with sensitivity and specificity derived from normalized counts of true positives, false positives, true negatives, and false negatives.

Baseline models for the lung, liver, and kidney analysis were constructed using only anthropometric measurements for comparison with those built using the combined anthropometric and EIT-derived features. ROC analysis assessing the diagnostic accuracy of these models was applied in the same way.

Because anthropometric measures have limited predictive value in heart failure, we constructed a more comprehensive cardiac baseline model using data from the UK Biobank. This baseline model incorporated demographic variables, blood pressure measurements, and multiple ECG-derived intervals and axes (e.g., QTc, QRS duration, RR/PP intervals, T/P axes). Importantly, because anthropometric and EIT cardiac measurements were obtained from different cohorts, the baseline model was derived solely from non-imaging features, whereas the EIT model was trained only on EIT-derived metrics.

### Statistical Analysis

Statistical analysis was performed using one of the follow tests: a one- or two-way analysis of variance, a linear mixed-effects model, a paired or unpaired (Welch’s) t-test, as needed for each experiment. A p-value < 0.05 was considered statistically significant. P-values were corrected for multiple comparisons when needed using the Benjamini-Hochberg false discovery rate approach. Specific test types, details, and p-values are provided in the main text and figure captions, and detailed statistical results are presented in Tables S1-8.

### Follow-up Questionnaires

To evaluate usability, participant satisfaction, and the feasibility of deploying VitoCheck at scale, we administered two optional post-screening questionnaires: one to participants and one to VitoCheck operators. Participants were invited to complete a brief survey documenting their experience with the VitoCheck test. Items included comfort during the scan, setup time, scan duration, and result wait time, clarity of explanation, perceived helpfulness of the health information provided, comparison to past clinical tests, likelihood of seeking medical follow-up, and willingness to undergo VitoCheck screening again.

Operators completed a separate survey evaluating ease of learning VitoCheck, confidence after first and subsequent uses, setup time, scan duration and result generation speed, perceived portability, feasibility for community-scale screening, and perceived value relative to typical imaging or laboratory tests.

## Data Availability

All data needed to evaluate the conclusions in the paper are present in the paper and/or the Supplementary Materials. The raw EIT data can be provided by RWC pending scientific review and a completed material transfer agreement. Requests for the raw EIT data should be submitted to. Sample datasets generated and analyzed during the current study will be made available in a public Zenodo/Dryad repository upon publication. Custom code used for EIT data analysis will be made available in a public GitHub repository upon publication.

## Acknowledgements

We thank Kannie W. Y. Chan, Eddie C. Wong, Way-Kay Seto, and Desmond Y.H. Yap for thoughtful insights and discussions. We also thank Vicky Huen, Lingfei Yin, Xiyu Chen, Kiyumi Lam, Vanie Lo, and Cheuk Man Ho for help with data acquisition.

## Funding

The work was funded in part by the Mitzi, Lee Leung Yin Ping Charitable Foundation, E-SENSE Innovation & Technology internal funds, and to a lesser extent the Hong Kong Innovation Technology Fund InnoHK Project at Centre for Cerebro-Cardiovascular Health Engineering

## Athor Contributions

Conceptualisation: WK, EF, RWC

Data Curation: JHWL, BJE, ML, RB

Formal Analysis: JHWL, BJE, ML, RB, RWC

Funding Acquisition: RWC

Investigation: JHWL, ML, RB, HL

Methodology: JHWL, BJE, WCK, ML, RB, HL, EF, RWC

Project Administration: WCK, KHY, RWC

Resources: WCKH, KHY, RWC

Software: JHWL, BJE, ML, RB, RWC

Supervision: RWC

Validation: BJE, WCK, ML, IYZ, KCC, KHY, EF, RWC

Visualisation: JHWL, BJE, ML, RB

Writing - Original Draft: JHWL, BJE, RWC

Writing - Review & Editing: JHWL, BJE, WCK, ML, RB, HL, IYZ, KCC, KHY, EF, RWC

## Competing Interests

RWC is a co-founder, shareholder and employee of E-SENSE Innovation & Technology Ltd., Hong Kong, China. BJE is a contractor and shareholder of E-SENSE Innovation & Technology Ltd., Hong Kong, China. JHWL is a consultant of E-SENSE Innovation & Technology Ltd., Hong Kong, China. The other authors do not have any conflict of interest.

